# Covid-19 disease dynamics with vaccination: The effect of uncertainty

**DOI:** 10.1101/2022.01.10.22269006

**Authors:** Abhijit Majumder, Nandadulal Bairagi

## Abstract

Rate parameters are critical in estimating the covid burden using mathematical models. In the Covid-19 mathematical models, these parameters are assumed to be constant. However, uncertainties in these rate parameters are almost inevitable. In this paper, we study a stochastic epidemic model of the SARS-CoV-2 virus infection in the presence of vaccination in which some parameters fluctuate around its average value. Our analysis shows that if the stochastic basic reproduction number (SBRN) of the system is greater than unity, then there is a stationary distribution, implying the long-time disease persistence. A sufficient condition for disease eradication is also prescribed for which the exposed class goes extinct, followed by the infected class. The disease eradication criterion may not hold if the rate of vaccine-induced immunity loss increases or/and the force of infection increases. Using the Indian Covid-19 data, we estimated the model parameters and showed the future disease progression in the presence of vaccination. The disease extinction time is estimated under various conditions. It is revealed that the mean extinction time is an increasing function of both the force of infection and immunity loss rate and shows the lognormal distribution. We point out that disease eradication might not be possible even at a higher vaccination rate if the vaccine-induced immunity loss rate is high. Our observation thus indicates the endemicity of the disease for the existing vaccine efficacy. The disease eradication is possible only with a higher vaccine efficacy or a reduced infection rate.

## I. INTRODUCTION

Vaccination against SARS-CoV-2 infection first started in the UK at the end of 2020 [1]. At present, ten WHO-recommended vaccine candidates are in use throughout the globe [2]. A good proportion of the population is vaccinated in the one year of its application [3]. Though the pandemic has not been controlled, the morbidity and mortality have been reduced significantly due to vaccination [4, 5]. Numerous mathematical models have been proposed and analyzed to determine the course of the covid pandemic since the WHO’s announcement of public health emergency of international concern (PHEIC) on January 30, 2020 [6] to restrict the spread of the novel coronavirus. These are mainly deterministic SEIR epidemic models or their variants [7–14] and a few are stochastic models [15–19]. Obviously, these earlier models did not consider the effect of vaccination. Unfortunately, such models can no longer be used for the epidemic course once the full-fledged Covid-19 vaccination has started. Therefore, any current time epidemic model should contain a vaccinated class. Recently, some researchers have proposed and analyzed some Covid-19 vaccination models to find the effect of immunization on the disease dynamics [20–22]. A Covid-19 vaccination model was studied in [20] to show that the waning of vaccine-induced immunity has a great impact on the disease dynamics. Without considering an explicit vaccination class, De la Sen and Ibeas [21] analyzed an SEIR type epidemic model to observe the combined role of vaccination and antiviral drugs in the controlling of COVID-19 pandemic. A case study of Japan shows that reduced vaccine efficacy and roll out of covid restriction may lead to a surge of covid cases. [22]. These are deterministic models and do not consider any uncertainty in the rate parameters. However, understanding the dynamics of a novel virus is insufficient if the inherent noise in the rate parameters is not considered. It is reported that there is uncertainty in the covid infection rate [23]. Due to spatial heterogeneity and other physical factors, there is significant variation in the Covid-19 recovery time, and rate [24, 25]. Most importantly, the efficacy of vaccines produced in an unprecedentedly shortest time is mostly unknown. It is also unclear how long these vaccines will provide protection against covid infection and to what extent. Even after taking a full dose of vaccine, it is now recommended for a buster dose, implying the loss of efficacy of vaccine [26, 27]. This indicates the existence of many uncertainties in the Covid-19 disease dynamics, its recovery rate, and vaccine efficacy. Therefore, it is essential to consider such uncertainties in the SARS-CoV-2 epidemic models with vaccination. Here we study a six-dimensional stochastic epidemic model to demonstrate the effect of vaccination in controlling the Covid-19 epidemic. Due to the variability in the infection rate, recovery rate, vaccine efficacy, we considered noise in these rate parameters and determined the disease persistence and eradication conditions. Using the Indian Covid-19 data, we estimated the best-fitted parameters and noise intensities. We then presented disease extinction time for the variation in the force of infection, vaccination rate, and immunity waning rate. Our analysis reveals that the Covid-19 disease will persist for a long time for the existing vaccine efficacy and transmissibility.

The remaining portion of this paper is organized in the following sequence. The stochastic Covid-19 vaccination model is proposed in the next Section II. Analytical results, including disease extinction conditions and stationary distribution of the solutions, are prescribed in Section III. Parameter estimation and the Indian case study are done in Section IV. The paper ends with a discussion in Section V.

## II. THE MODEL

We propose an extended SEIR stochastic compartmental epidemic model to investigate the Covid-19 disease under vaccination. The total human population, *N*(*t*), of a region is divided into six mutually exclusive groups, viz., susceptible, exposed, detected infectives, undetected infectives, recovered, and vaccinated, which are denoted by *S, E, I, A, R,V*, respectively. The susceptible individuals are recruited through birth at a rate Λ. After coming into the effective contact of a detected or undetected covid infected individual, susceptible individuals become infected and join the *E* class, who carry the virus but are not yet infectious. The transmission probability of covid infection from the detected and undetected individuals may differ. Assume that *κ* be the transmission probability of infection due to the contact between susceptible and infected individuals. The same is (1 −*κ*) for the contact between susceptible and detected individuals. If *β* be the average per capita daily contact, then the susceptible individual that joins the *E* class is given by 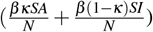. Since the vaccination of a susceptible individual does not give 100 % immunity against coronavirus, the vaccinated people may again be infected from the undetected and detected individuals, but possibly at a lower rate. Considering *η* (*η < β*) as the disease transmission efficiency, the portion of the vaccinated individuals who join the exposed class at time *t* is 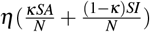. It is observable that *η* measures the vaccine efficacy. If *η* = 0, then the vaccine will be 100% effective. An exposed class individual spent on an average 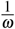 time in *E* class, and then joined either the undetected class with probability *δ* or the detected class with probability (1 − *δ*). An average time of 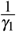 and 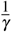 are spent by the undetected and detected individuals, respectively, before moving to the recovered class. Recovered people can also lose immunity and join the susceptible class at a rate of *g*. Susceptible people are vaccinated at a rate *q* and join the *V* class. Natural death at a rate *m* is incorporated in every compartment, and an additional disease-related death rate *d*_*i*_ is included in the detected class, *I*. We do not consider any disease-related death in the *A* class because critically ill individuals, if any, may be shifted to the *I* class at a rate *ν*. Since the coronavirus is a novel virus, there are substantial uncertainties in the rate constants, like infection rate [28], recovery rate [29] in *A* and *I* classes, and also in the rate of immunity loss [30]. To incorporate this uncertainty, we gave random perturbations to these parameters as follows:

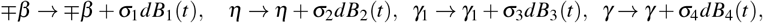

where *B*_*i*_(*t*) are standard mutually independent Brownian motions and 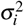, *i* = 1,2,3,4 are the intensities of the white noises. Similar parametric perturbation has also been considered in other biological models [31–35]. Encapsulating all these assumptions, the stochastic compartmental model for the Covid-19 vaccinated model reads

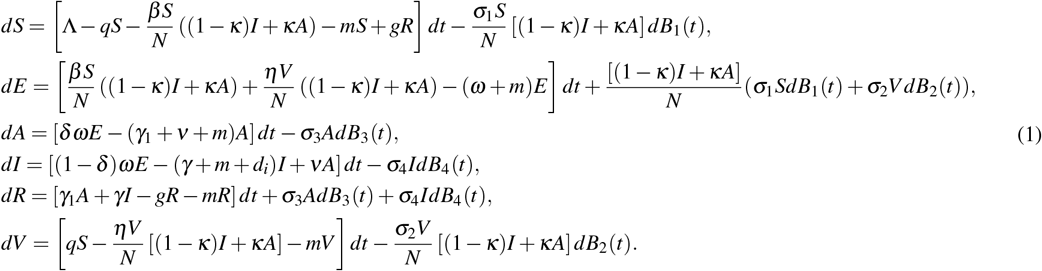

The initial values for the state variables are considered as

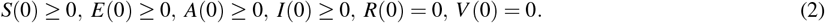

## III. RESULTS

It is imperative to show that the considered system has a unique global solution, and all the solutions are positive when starting with positive initial values. We have the following theorem for this result.

### Theorem III.1

For any initial value 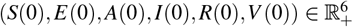, there exists a unique solution of the system (1) such that 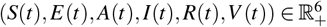 for all *t* ≥ 0 and the solution remains in 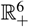 with probability 1, i.e., almost surely (a.s).

*Proof*. The proof is given in the Appendix I. □

It is worth mentioning that the stochastic system (1) has no equilibrium point. However, it may have some stationary distribution, meaning that the asymptotic solution of the system does not change significantly. From an epidemic point of view, it implies the long-term persistence of the disease. We show that such stationary distribution occurs if the following theorem holds good.

### Theorem III.2

Assume that

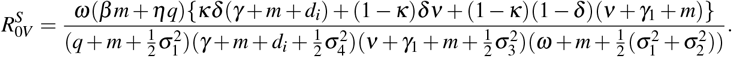

Then, for any initial value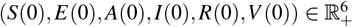, a sufficient condition for existing a stationary distribution *π*(.) of the system (1) is 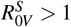.

*Proof*. Please see Appendix-II for its proof. □

Here 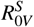 may be called as the stochastic basic reproduction number (SBRN), which ensures the disease establishment in the stochastic system (1). One can easily obtain the deterministic basic reproduction number (DBRN), which is the disease existence condition in a deterministic epidemic model, as 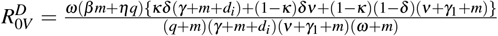 (see Appendix - III). Note that if *σ*_*i*_ = 0, *i* = 1,.., 4, then 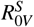 and 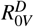 are equivalent. Also, the basic reproduction number of the stochastic system 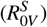 is smaller than that of the corresponding deterministic system 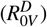.

Observe that both the infected classes (symptomatic and asymptomatic) originate from the exposed class. Thus, the infection will eventually be eradicated if the individuals of the exposed class go extinct. For the system (1), the exposed class *E*(*t*) is said to be extinct (i.e., the system will be disease-free) if lim_*t*→∞_ *E*(*t*) = 0 *a*.*s*. [15]. We give here some sufficient conditions for which the exposed class dies out over time. In proving the extinction criterion, the result of strong law of large number given in the following lemma will be used.

### Lemma III.1

[36] Let *M* = {*M*}_*t*≥0_ be a continuous valued local martingale and vanishing at *t* = 0, then

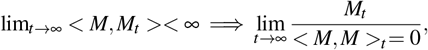

and

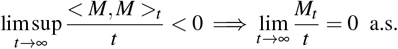

### Theorem III.3

The exposed individuals of the system (1) tend to zero exponentially almost surely if 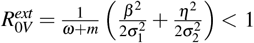.

*Proof*. Assume that 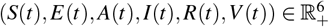 is a solution of system (1) satisfying the initial value 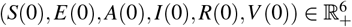. Following Ito’s formula, we have from (1)b

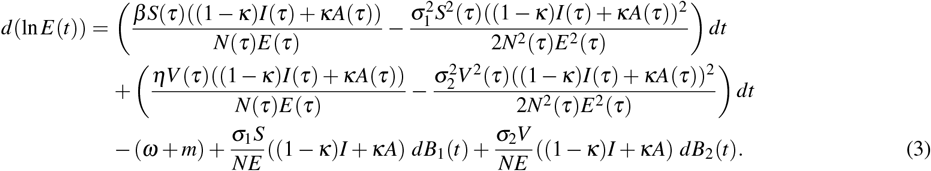

Upon integration from 0 to *t*, we have

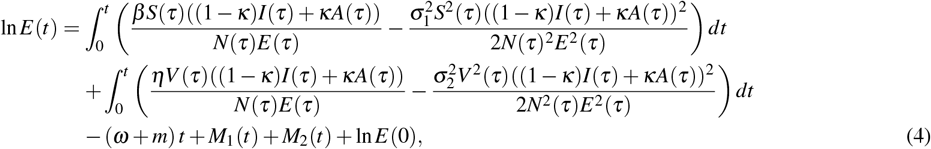

where

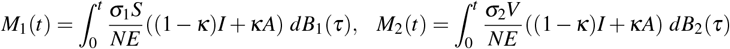

are the local continuous martingale with *M*_1_(0) = 0, *M*_2_(0) = 0. We, then have

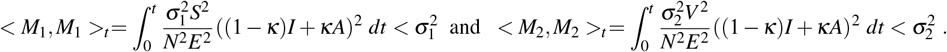

Using the fact

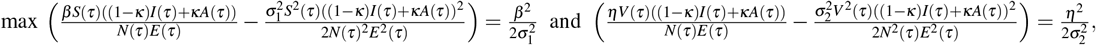

(4) can be written as

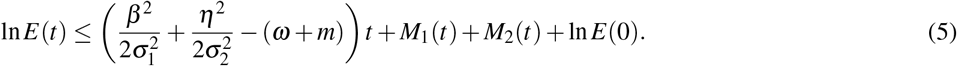

Taking the limit superior as *t* → ∞, after dividing both sides of (5) by *t* (> 0) and using Lemma III.1, we have

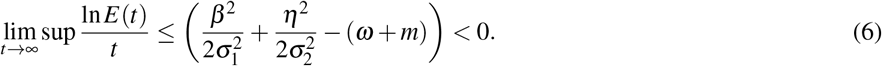

If 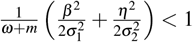, then lim _*t*→∞_ *E*(*t*) = 0 almost surely. Hence the theorem. □

It is observable that 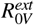 is an increasing function of *β* and *η*. Thus, if the infection rate increases or the vaccine-induced immunity loss increases, the inequality 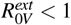 may not be held, and consequently, the disease eradication may not be possible.

## IV. CASE STUDY

For the case study, we considered the Indian Covid-19 epidemic data available from the repositories Covid19India.Org (https://covid19india.org) and Worldometers.info (https://www.worldometers.info/coronavirus/country/india/). The daily and cumulative numbers of infected, recovered, deceased, and vaccinated cases are reported and updated daily in these depositories. We had considered only the Covid data when the second wave started in India. The daily infected cases of the first wave reached their minimum value (8579 cases per day) on 1^*st*^ February 2021 (https://covid19india.org). Therefore, the 2^*nd*^ February 2021 was considered as the starting date of the second wave, and we considered the data from this date to 31^*st*^ December 2021 for parameter estimation. For better fitting of the curve, the whole data set was divided into two intervals: from 2^*nd*^ February to 6^*th*^ May (the date when the peak is attained in the second wave) and from 7^*th*^ May to 31^*st*^ December 2021. Indian Covid-19 vaccination program started on January 16, 2021 [37]. Though the initial vaccination rate was slow, it was intensified later on. We fitted (see Fig. 1) the actual covid data (in red color) for the considered period with the stochastic model solution (in blue color). The best-fitting parameter values and the optimal noise intensities are provided in Table I. The parameters and noise estimation technique are given in Appendix-IV.

**TABLE I.** Estimated parameter values for India

| Period | $\Lambda$ | $m$ | $\beta$ | $\kappa$ | $\delta$ | $\omega$ | $\gamma$ | $\gamma_1$ | $g$ | $d_i$ | $v$ | $\eta$ | $q$ | $\sigma_1$ | $\sigma_2$ | $\sigma_3$ | $\sigma_4$ |
| --- | --- | --- | --- | --- | --- | --- | --- | --- | --- | --- | --- | --- | --- | --- | --- | --- | --- |
| 2<sup>nd</sup> February to 6<sup>th</sup> May, 2021 | 77756 | $4.1 \times 10^{-5}$ | 0.115 | 0.92 | 0.66 | 0.18 | 0.03 | 0.032 | 0.004 | 0.007 | 0.002 | 0.11 | $1.6 \times 10^{-3}$ | 0.03 | 0.02 | 0.035 | 0.04 |
| 7<sup>th</sup> May to 31<sup>st</sup> December, 2021 | 77756 | $4.1 \times 10^{-5}$ | 0.109 | 0.92 | 0.66 | 0.18 | 0.014 | 0.16 | 0.064 | 0.018 | 0.002 | 0.23 | $2.1 \times 10^{-3}$ | 0.06 | 0.05 | 0.04 | 0.05 |

**FIG. 1.**
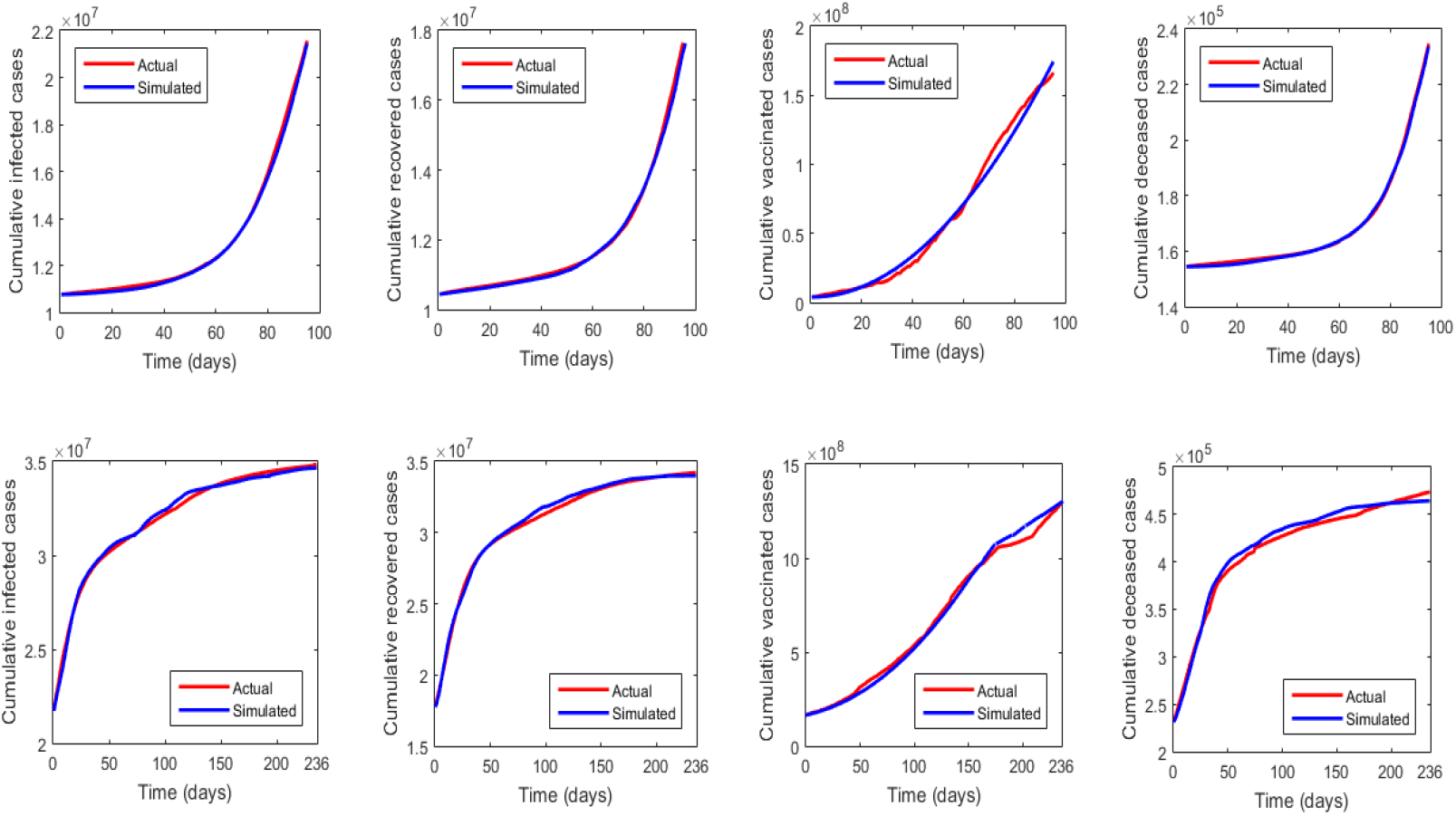
Covid-19 data fitting. Upper row: The cumulative actual covid data (magenta color curve) of the confirmed, recovered, vaccinated and death cases in India for the study period February 2 to May 6, 2021, are fitted by the solution (blue color curve) of the stochastic model (1). Noise intensities are *σ*_1_ = 0.03, *σ*_2_ = 0.02, *σ*_3_ = 0.035, *σ*_4_ = 0.04. Lower row: The same is plotted for the period May 7 to December 31, 2021 in the bottom row. Here noise intensities are *σ*_1_ = 0.06, *σ*_2_ = 0.05, *σ*_3_ = 0.04, *σ*_4_ = 0.05.

We then investigate the mean disease eradication time if the vaccination-induced immunity-loss parameter, *η*, is varied. Figure 2 (upper left) shows that the disease may be eradicated within the next 275 days if the vaccine provides 100% protection from the reinfection. The extinction time gradually increases if the vaccine-induced immunity loss increases. The mean extinction time is drastically increased, and the extinction curve becomes an asymptote once this loss is above 25% (i.e., *η* > 0.25), implying that the disease eradication is virtually impossible. The corresponding probability density curve (Figure 2, upper right) shows the lognormal distribution of mean disease eradication time. It indicates that the disease eradication time over 700 days is almost nil. A similar estimation of the disease eradication time (Figure 2, lower left) is also observed for the variation of infection rate (*β*). At the very low infection rate, the disease extinction time is approximately 230 days, and the required time increases with increasing transmissibility when other parameters remain unchanged. The probability density function, in this case, is also best fitted by a lognormal distribution (Figure 2, lower right).

**FIG. 2.**
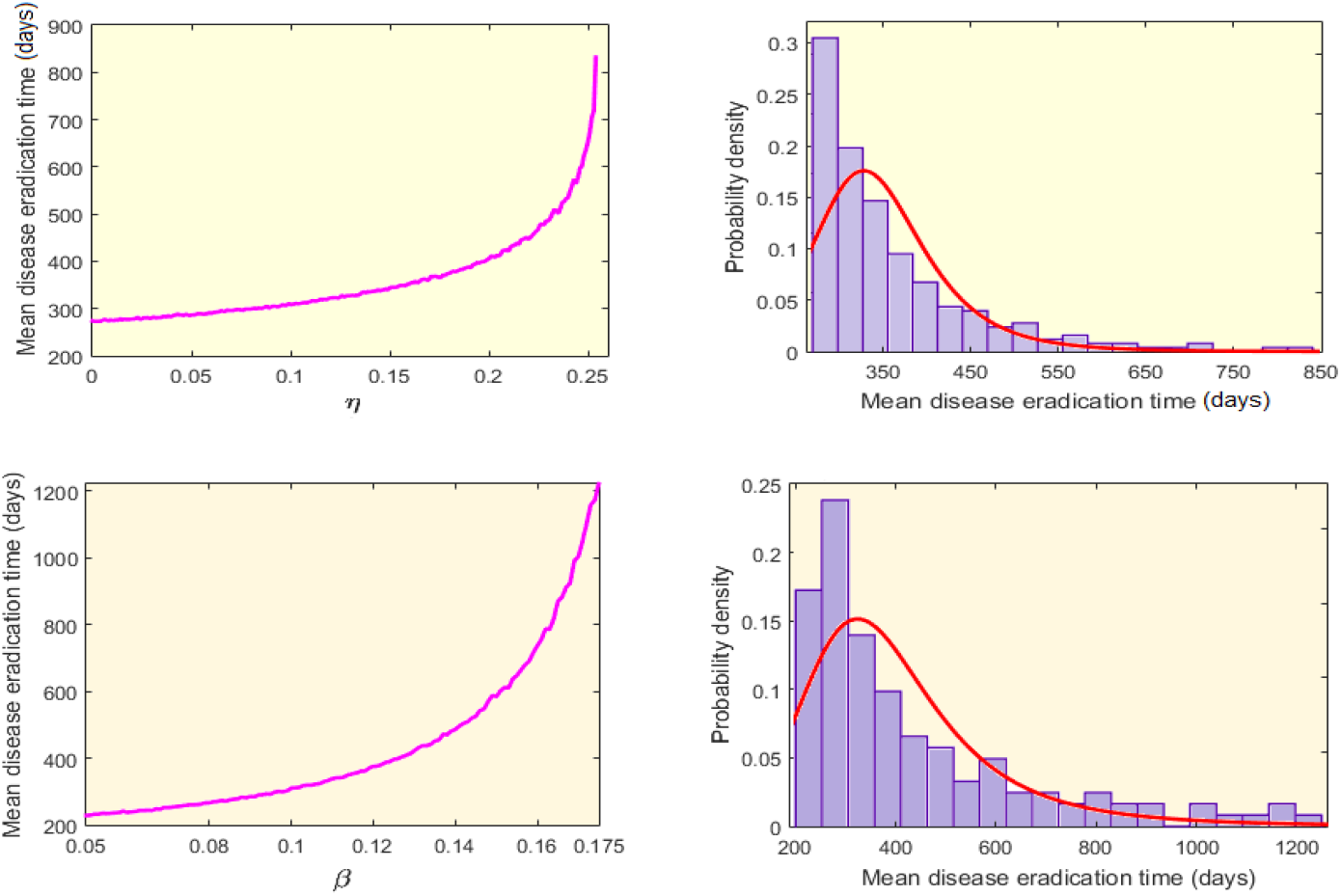
Mean disease eradication time of the stochastic model (1). Upper row: For the variation in the parameter *η* (left figure), and the corresponding probability density curve (right-hand figure). Lower row: with respect to *β* (bottom left) and the corresponding probability density curve (bottom right). Noise intensities and parameter values are for the period May 7 to December 31, 2021, see Table I.

We now examine how the vaccination rate (*q*) and the immunity loss rate (*η*) influence the disease burden. Figure 3 represents that the per day covid positive cases increases with increasing vaccination rate if vaccine-induced immunity loss exceeds the value 0.24, i.e., the vaccine efficacy is lower than 76%. Observe that the per day cases becomes as high as 1.7 million when *η* = 0.26 and *q* = 0.22. Thus, increased vaccination cannot mitigate covid infection under low vaccine efficacy, and rather it increases the covid cases. However, infection eradication is possible for the considered range of vaccination rate if the vaccine’s immunity is more than 76%. It is to be mentioned that 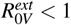 holds for most of the lower values of *q* and *η* and 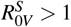 for its higher values.

**FIG. 3.**
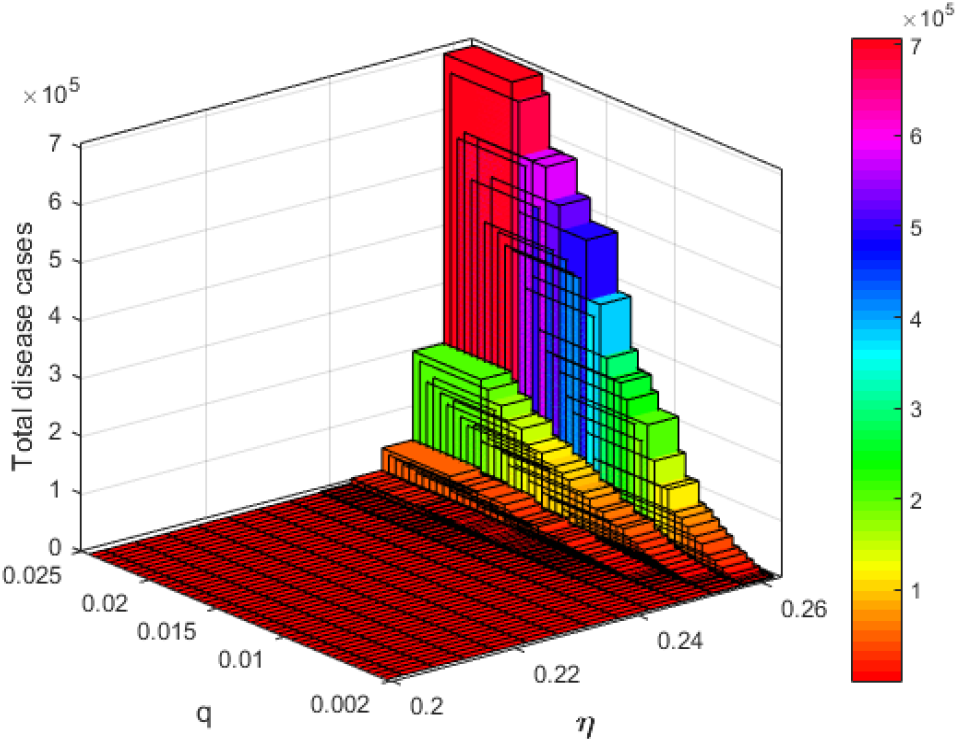
Per day covid positive cases for the variable rates of *η* and *q*. The end average value (asymptotic plus symptomatic) of 1000 simulations for 1000 time steps for each value of *η* and *q* is plotted here. Noise intensities and other parameter values remain fixed as in the period May 7 to December 31, 2021, see Table I.

A similar phenomenon is plotted in Fig. 4 when the mean infected cases per day are plotted for the variable vaccination rate and force of infection (*β*). The covid positive cases gradually increase if *β* is high and *q* is low. The number of positive cases may be 0.2 million per day at a low vaccination rate and high transmission (left figure). In the opposite case, the disease, however, may be eradicated. The right-hand figure represents the newly infected per day covid cases when the force of infection (*β*) and the vaccine effectivity (*η*) parameters are jointly varied. The infection spreads rapidly in the parametric space, where *β* > 0.2 and *η* > 0.23 (Fig. 4, right). The covid case may be as high as 1.4 million per day in this parametric range. It demonstrates that vaccine effectiveness plays a more crucial role in the new cases.

**FIG. 4.**
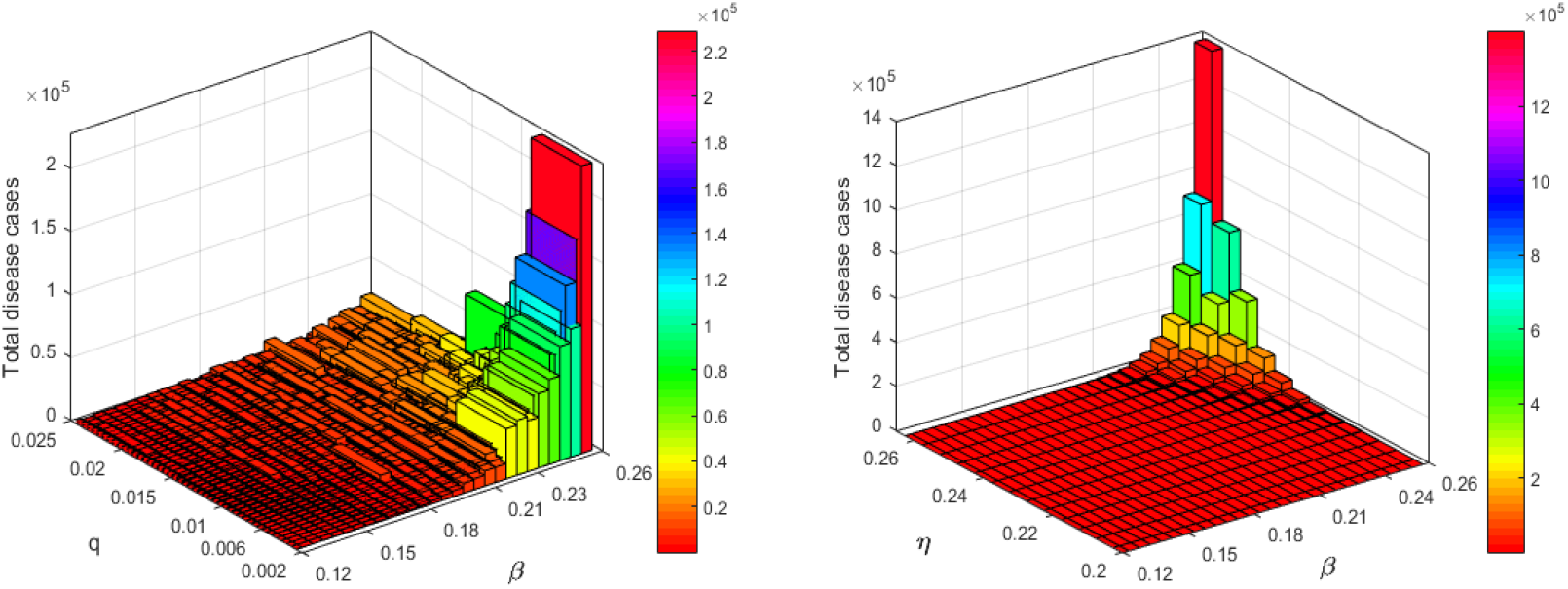
Per day covid positive cases with respect to *β* and *q* (left) and *β* and *η* (right). Noise intensities and other parameter values remain fixed as in the period May 7 to December 31, 2021, see Table I.

We further explored the effect of noises in the total number of covid cases for the default parameter values (see Figure 5). On the left of this figure, cumulative covid cases are plotted for four different noise intensities of *σ*_1_ with a 95% confidence interval. A similar figure is drawn for the noise *σ*_2_. It is to be observed that the noise intensity *σ*_2_, which is attached to the parameter *η*, has a higher impact on the covid cases. It is worth mentioning that the other two noises have a much lower effect compared to the first two (figure not provided).

**FIG. 5.**
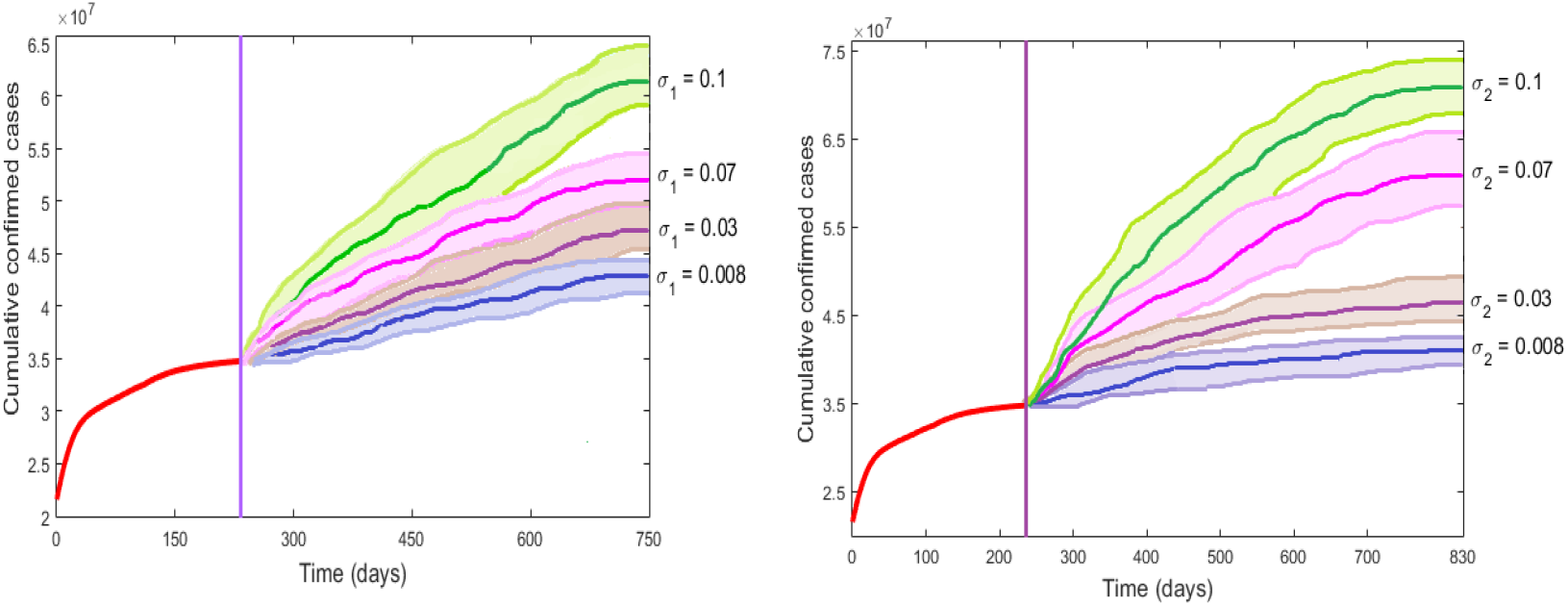
The cumulative covid case (*A* + *I*) with 95% confidence interval for different noise intensities of *σ*_1_ (left figure) and *σ*_2_ (left figure). Parameters remain fixed as in the period May 7 to December 31, 2021, see Table I.

## V. DISCUSSION

The SARS-CoV-2 infection has kept the world under pressure for two years. Most countries have experienced more than two waves of this infection at the cost of millions of lives. Massive vaccination program started at the end of 2020, hoping that the disease will be controlled. Though the morbidity and mortality of the covid disease reduced significantly, the disease eradication even of its control is far from expected. Many countries have been experiencing higher positive cases in the ongoing wave than the previous peak values. Several European countries, the UK, the USA have fully vaccinated a significant proportion of their population but cannot resist further covid infection. This fact has put the efficacy of the vaccine under question. Recent studies show that vaccine-induced immunity is significantly reduced after six to eight months post-vaccination. The level of a covid antibody that persists after this period may not be sufficient to prevent reinfection. There is uncertainty regarding the rate of immunity loss among the vaccinated population. Other uncertainties exist in different rate parameters, e.g., the force of infection and recovery rates. It is undoubtedly true that the omicron variant’s infectivity is much higher than in previous strains. Also, the severity of the disease is relatively low, and the recovery rate is significantly elevated in the current wave caused due to the omicron variant of coronavirus. Considering such uncertainties in the rate parameters, we have proposed and analyzed a six-dimensional stochastic Covid-19 epidemic model in the presence of vaccination. The objective was to understand the effect of vaccination on the disease dynamics in the presence of noise. We here prescribed both the disease persistence and eradication conditions. It is shown that the disease persists for a long time almost surely if the stochastic basic reproduction number (SBRN) is greater than unity. It is noticed that this value of SBRN is smaller than the DBRN (deterministic basic reproduction number) of the corresponding deterministic model, which is usually used as a measure of disease establishment in the latter type of epidemic models. We have also proved a sufficient condition for disease eradication. It is shown that the disease cannot persist if 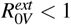. Noticeably, this condition may not hold if the disease’s infectivity increases or/and the vaccine-induced immunity loss increases. Both issues are probably real for many countries, where vaccination started in the initial months of 2021, and the new variant, omicron, is responsible for the ongoing wave. We used the Indian Covid-19 data to estimate the model parameters and the noise intensities. We have demonstrated the disease extinction time under different scenarios using the estimated parameter values. It is revealed that the mean extinction time increases with the increasing rate of immunity loss and force of infection. The probability density curves for the parameters show the lognormal distribution of mean disease eradication time. Noticeably, if the vaccine-induced immunity loss rate, *η*, is higher than 0.24, eradication of infection is practically impossible. The case is almost similar if the infectivity is also high. It implies that the disease will last long unless a long-lasting vaccine candidate appears or a low infectious variant replaces the highly infectious variant.

## Data Availability

Covid19India.Org and Worldometers.info

https://covid19india.org

https://www.worldometers.info/coronavirus/country/india/

## Acknowledgment

Research of Abhijit Majumder is supported by CSIR (File No: 09/096(0874)/2017-EMR-I)

## Appendices

### Appendix I

Since the coefficients of the model system (1) are locally Lipschitz continuous, for any 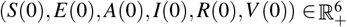, there is a unique local solution 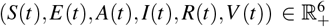 for all *t* ∈ [0, *τ*_*e*_), where *τ*_*e*_ is the explosion time [38]. We now prove *τ*_*e*_ = ∞ a.s. so that the solution becomes global. Let *κ*_0_ > 0 be sufficiently large for every coordinate (*S*(0),*E*(0),*A*(0), *I*(0),*R*(0),*V*(0)) lying within the interval 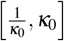. We then define, for every integer *κ* > *κ*_0_, the stopping time

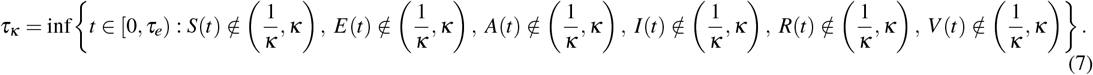

Thus, *τ*_*κ*_ is increasing as *κ* → ∞. Set lim_*κ*→∞_ *τ*_*κ*_ = *τ*_∞_, when *τ*_∞_ ≤ *τ*_*e*_ a.s. We now show that *τ*_∞_ = ∞ by a contradiction. Let us assume that our claim is not true and there exist two constants *T* > 0 and *ε* ∈ (0, 1) such that

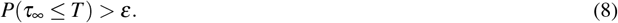

Thus, there exists an integer *κ*_1_ ≥ *κ*_0_ such that

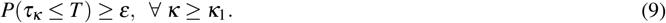

Noticing that *u* + 1 − ln *u* > 0 for all *u* > 0 and 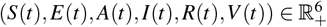, we define the following positive definite function

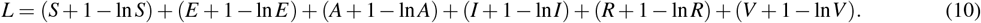

Applying Ito’s formula, one can have

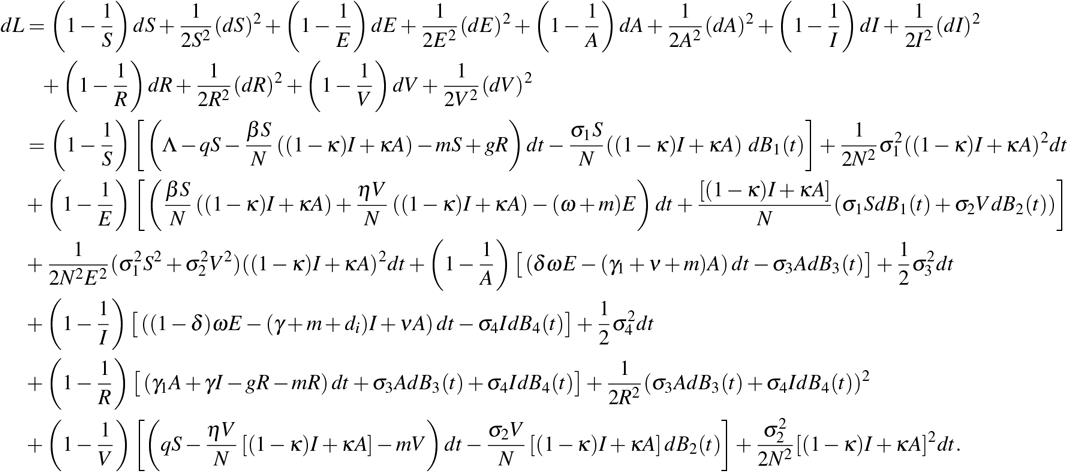

Noting *u* ≤ 2(*u* + 1 − ln *u*) for all *u* > 0 and *N* is the total population, the above expression becomes

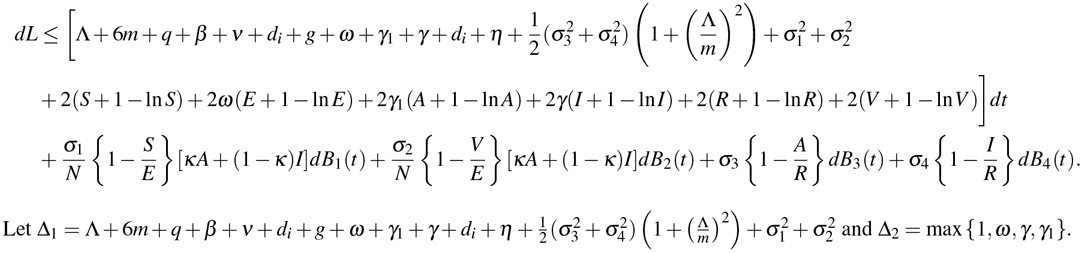

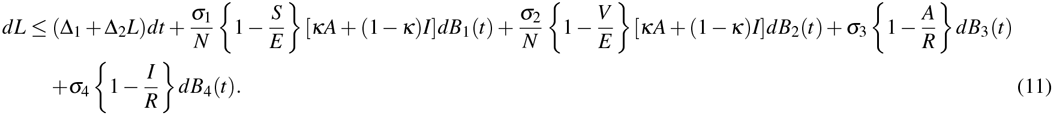

Defining Δ_3_ = max{Δ_1_, Δ_2_}, we have

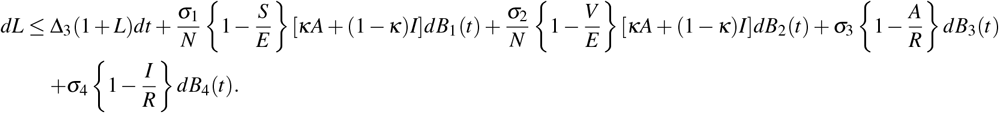

Noticing that

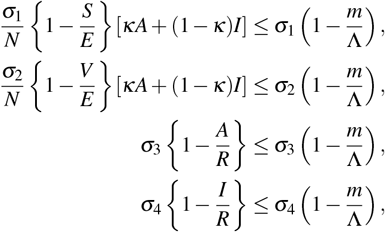

we have

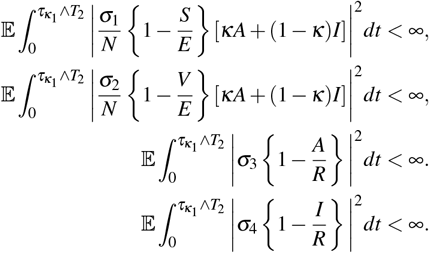

Since all the functions 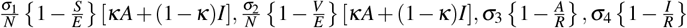 are continuous, bounded and non-anticipative, then for a sequence of partition of the interval 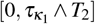 with mesh size Δ*t* → 0, we have

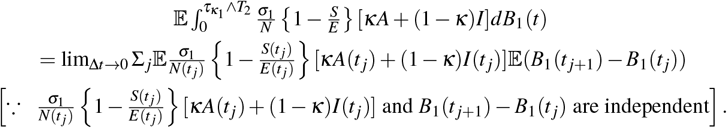

Similarly, we have

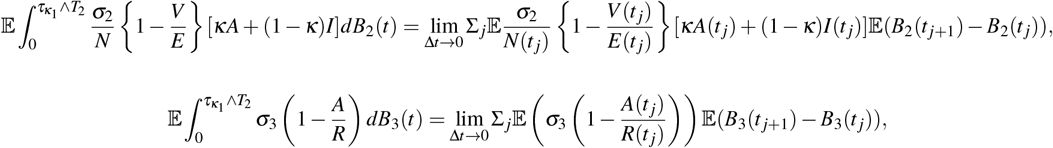

and

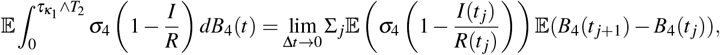

Using the fact that the increment of the Brownian motion are normally distributed with mean zero and variance (*t* _*j*+1_ − *t* _*j*_), we have

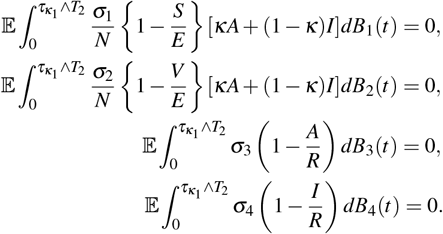

Integrating both sides of (12) from 0 to 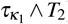, taking the expectation and using the above fact, we obtain

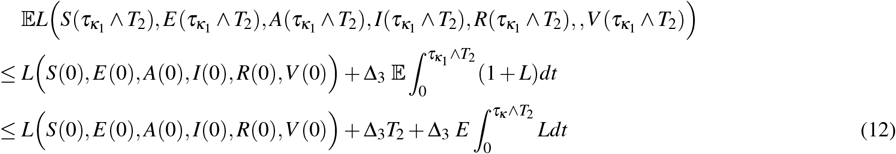

Since *L* is an increasing function on 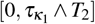, hence for any *t* ∈ 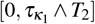,

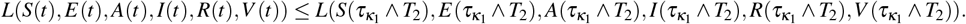

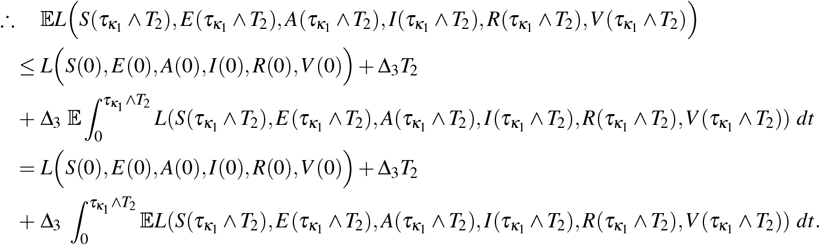

Gronwall’s inequality then gives

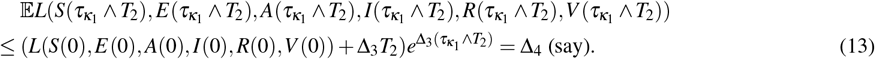

Set 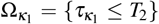 for all *κ*_1_ ≥ *κ*_2_. Thus, following (9), we get 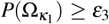 for all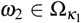. Clearly, at least one of 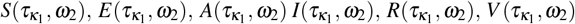 is equal to either *κ*_1_ or 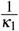. Hence, 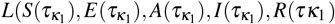, is no less than min 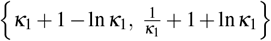. From (8) and (13), we then obtain

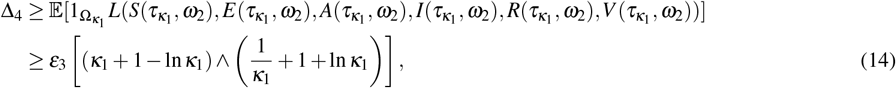

where 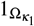 is the indicator function of 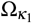. Letting *κ*_1_ →∞, we get ∞ > Δ_4_ = ∞, a contradiction. Hence *τ*_∞_ = ∞ a.s. Hence the theorem is proven.

### Appendix II

In order to show the existence of a stationary distribution, we here adopted the technique given in [40]. The following lemma will be used in the sequel.

#### Lemma VI.1

[39] Let *X*(*t*) be a regular Markov process (time-homogeneous) in 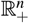 whose dynamics is described by the stochastic equation

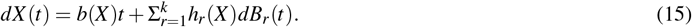

Then the corresponding diffusion matrix is defined as

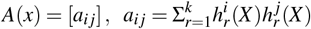

and the solution *X* (*t*) of (15) has a unique stationary distribution *π*(.) if there exist a bounded domain *U* ∈ *R*^*n*^ with regular boundary Γ and (a) there is a positive number *M*_2_ such that 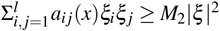, (b) there exist a nonnegative *C*^2^-function *V*_1_ such that *LV*_1_ is negative for any *R*^*d*^\*U*. Then

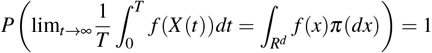

for all *x* ∈ *R*^*d*^, where *f* (.) is a function integrable with respect to the measure *π*.

By Theorem III.1, for any initial size of population 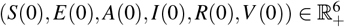, there exists a unique non-local global solution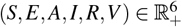. Let us denote 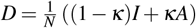. The diffusion matrix of the system (1) is given by

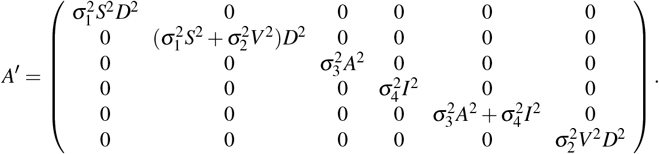

Let 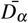 is a bounded domain in 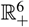 which excludes the origin. Choose 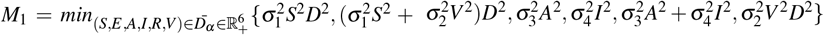. For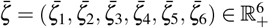, we can obtain

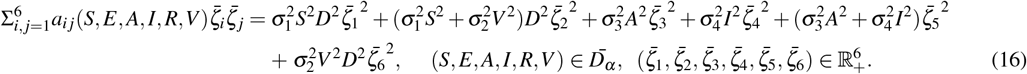

Hence the condition (a) of Lemma VI.1 holds. In order to prove the second assertion of the lemma, define a non-negative *C*^2^ function *H*_1_, where *H*_1_: 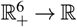 R be such that

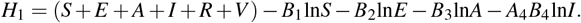

Applying Ito formula, one gets

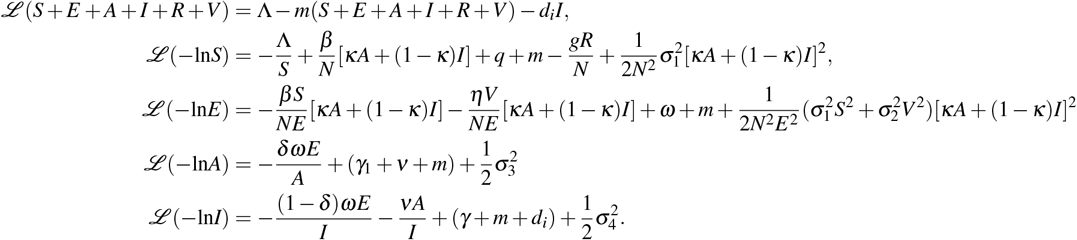

Therefore, we have

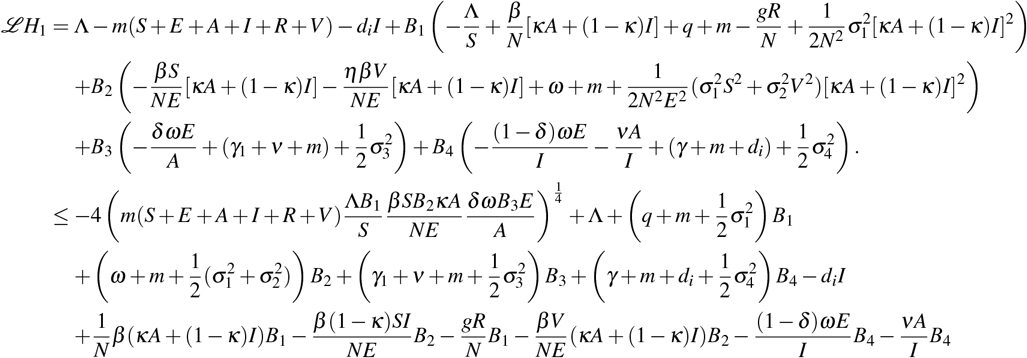

Define 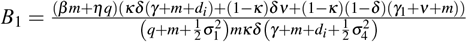 and let

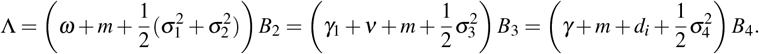

Therefore,

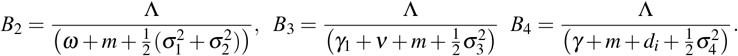

Consequently,

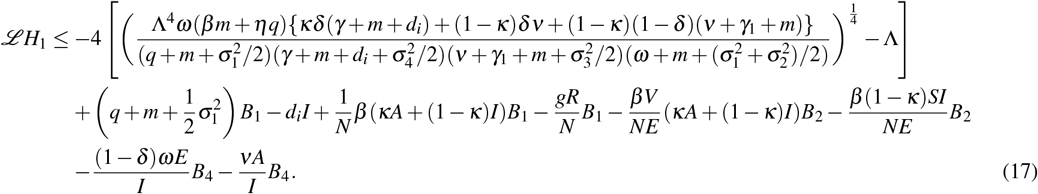

Define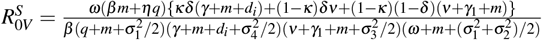, so that (17) becomes

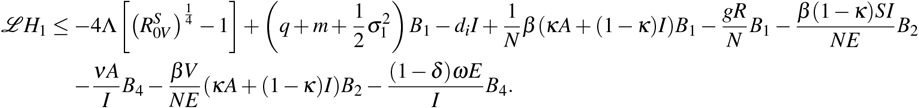

We further define

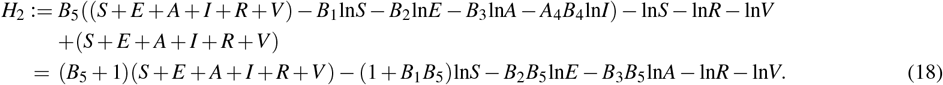

Let 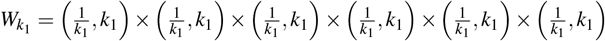. As *k*_1_ → ∞, it is evident that

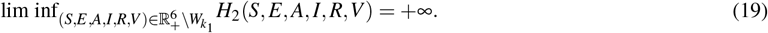

Now, we intended to prove that *H*_2_(*S, E, A, I, R,V*) has the unique smallest value *H*_2_(*S*(0), *E*(0), *A*(0), *I*(0), *R*(0),*V* (0)). Taking partial derivatives of the function *H*_2_(*S, E, A, I, R,V*) with respect to each state variable, we get

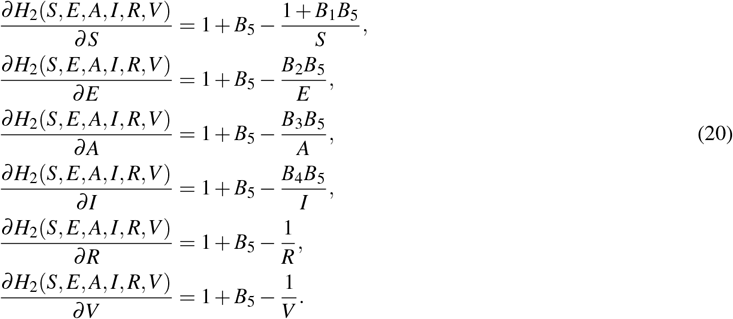

Making each of these partial derivatives equal to zero, we get 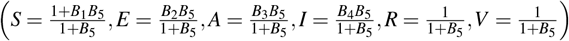 as the unique stagnation point of *H*_2_. Furthermore, the Hesse matrix of the function *H*_2_(*S, E, A, I, R,V*) at the given initial population density reads

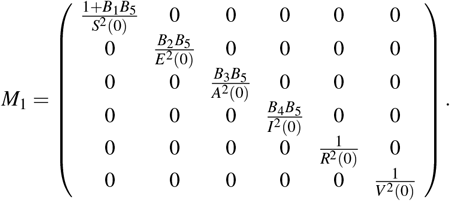

Clearly, the matrix *M*_1_ is positive definite. Hence *H*_2_ attains the smallest value at 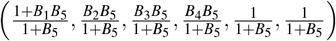. From the continuity of *H*_2_ and using equation (19), we can state that inside 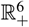, the function *H*_2_(*S, E, A, I, R,V*) has the unique smallest value *H*_2_(*S*(0), *E*(0), *A*(0), *I*(0), *R*(0),*V* (0)). We, now define a non-negative *C*^2^ function *H*: 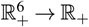 such that

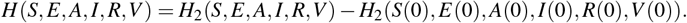

Applying Ito formula on *H* and using the model (1), one can obtain

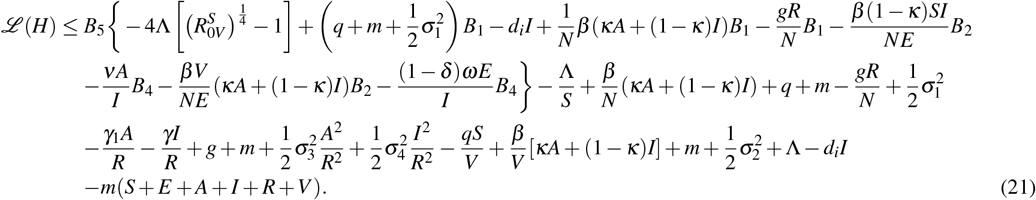

Under the assumption 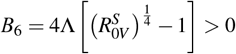, (21) becomes

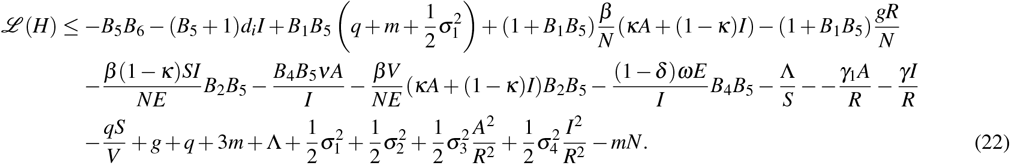

Consider now the following bounded subset

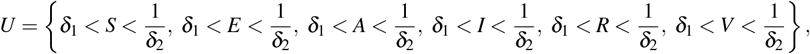

where *δ*_*i*_ > 0 for *i* = 1, 2 are negligibly small constants to be chosen later on. We divide the domain 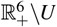 into the following sub-domains:

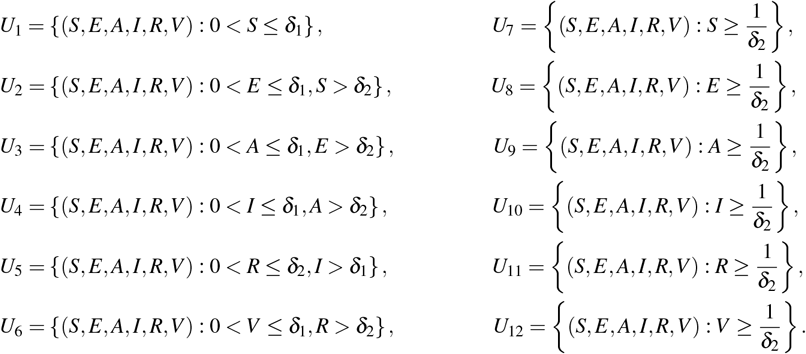

We will now show that *ℒH*(*S, E, A, I, R,V*) *<* 0 on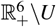, or equivalently, *ℒH <* 0 in all of the above twelve regions. We will discuss *ℒH* in each region one by one.

**Case** 1. Suppose (*S, E, A, I, R,V*) ∈ *U*_1_, then (22) becomes

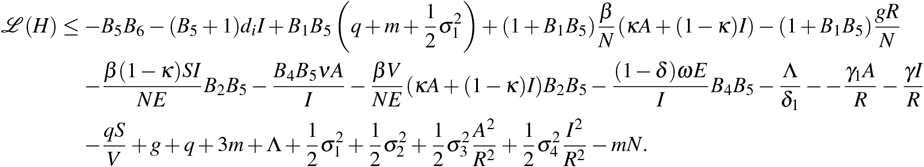

Choosing *δ*_1_ > 0 sufficiently small, one obtains *ℒ*(*H*) *<* 0 for every (*S, E, A, I, R,V*) ∈ *U*_1_.

**Case** 2. If (*S, E, A, I, R,V*) ∈ *U*_2_, then from (22), we can obtain

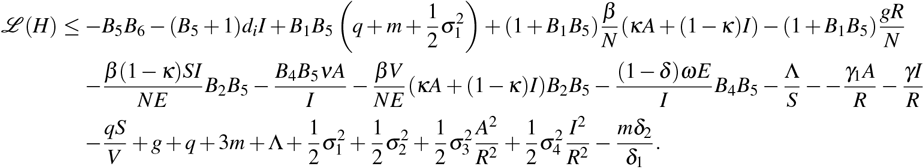

Let 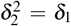 and choosing large positive value of *B*_5_ and sufficiently small value of *δ*_2_, one have *ℒ*(*H*) *<* 0 for every (*S, E, A, I, R,V*) ∈ *U*_2_.

**Case** 3. Assuming (*S, E, A, I, R,V*) ∈ *U*_3_, one obtains from (22),

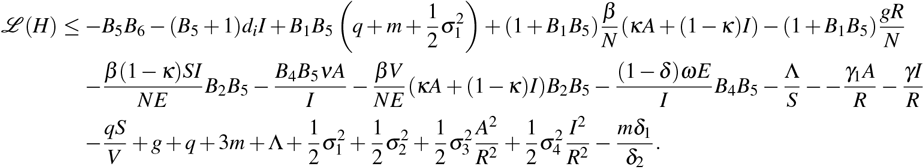

Let 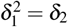 and selecting sufficiently small value of *δ*_1_, we can get *ℒ*(*H*) *<* 0 for every (*S, E, A, I, R,V*) ∈ *U*_3_.

**Case** 4. If (*S, E, A, I, R,V*) ∈ *U*_4_, equation (22) yields

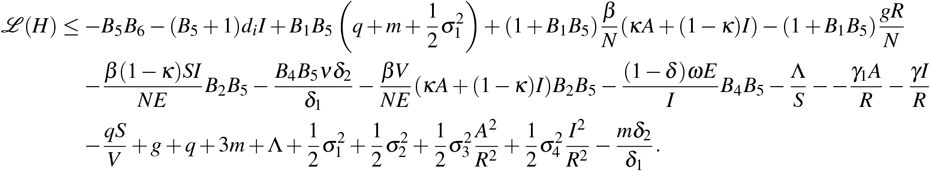

Assuming 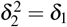 and choosing sufficiently small value of *δ*_2_, we can get *ℒ*(*H*) *<* 0 for every (*S, E, A, I, R,V*) ∈ *U*_4_.

**Case** 5. If (*S, E, A, I, R,V*) ∈ *U*_5_, from (22), one can obtain

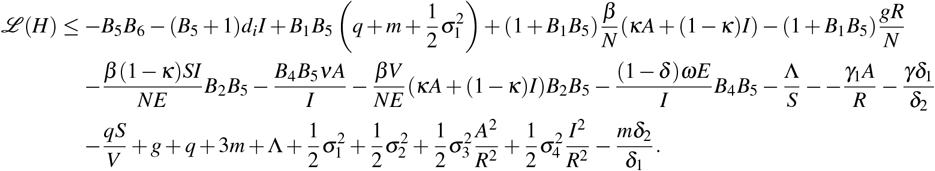

Assuming 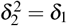 and choosing sufficiently small value of *δ*_2_, we can get *ℒ*(*H*) *<* 0 for every (*S, E, A, I, R,V*) ∈ *U*_5_.

**Case** 6. If (*S, E, A, I, R,V*) ∈ *U*_6_, from (22), one can obtain

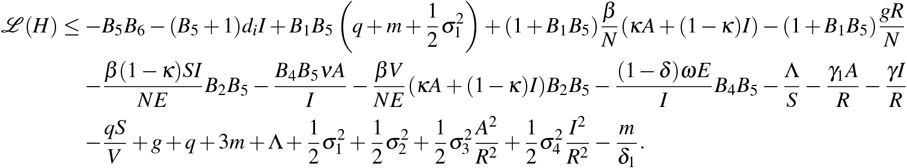

Choosing sufficiently small value of *δ*_1_ > 0, we can get *ℒ*(*H*) *<* 0 for every (*S, E, A, I, R,V*) ∈ *U*_6_.

**Case** 7. If (*S, E, A, I, R,V*) ∈ *U*_7_, from equation (22), one can obtain

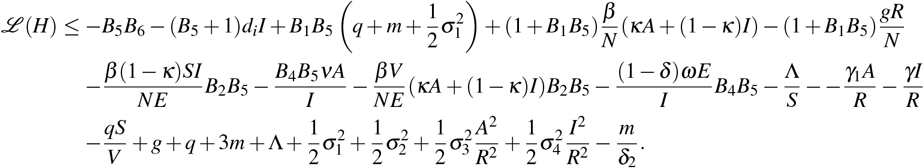

In order to make *ℒ*(*H*) *<* 0 for every (*S, E, A, I, R,V*) ∈ *U*_7_, we choose sufficiently small value of *δ*_2_ > 0.

**Case** 8. If (*S, E, A, I, R,V*) ∈ *U*_8_, from equation (22), one can obtain

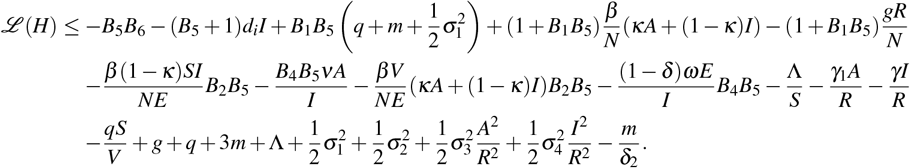

Considering sufficiently small value of *δ*_2_ > 0, we can find *ℒ*(*H*) *<* 0 for every (*S, E, A, I, R,V*) ∈ *U*_8_.

**Case** 9. If (*S, E, A, I, R,V*) ∈ *U*_9_, from equation (22), one can obtain

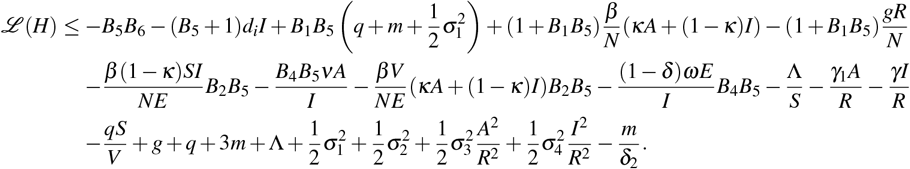

Upon choosing sufficiently small value of *δ*_2_ > 0, we have *ℒ*(*H*) *<* 0 for every (*S, E, A, I, R,V*) ∈ *U*_9_.

**Case** 10. Consider the case when (*S, E, A, I, R,V*) ∈ *U*_10_. Then equation (22) yields

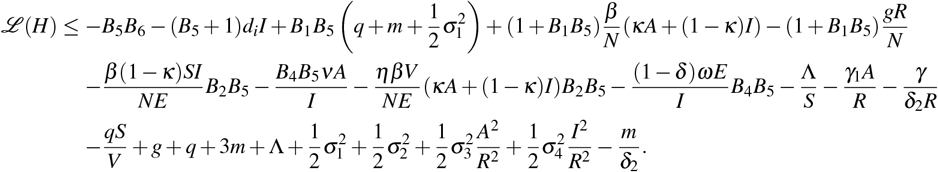

By choosing sufficiently small value of *δ*_2_ > 0, we have *ℒ*(*H*) *<* 0 for every (*S, E, A, I, R,V*) ∈ *U*_10_.

**Case** 11. In the case when (*S, E, A, I, R,V*) ∈ *U*_11_, equation (22) reduces to

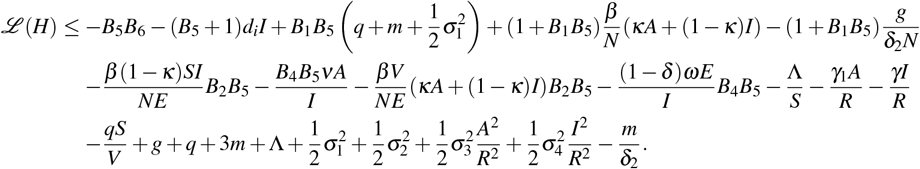

For sufficiently small value of *δ*_2_ > 0, one can obtain *ℒ*(*H*) *<* 0 for every (*S, E, A, I, R,V*) ∈ *U*_11_.

**Case** 12. Upon choosing (*S, E, A, I, R,V*) ∈ *U*_12_, equation (22) becomes

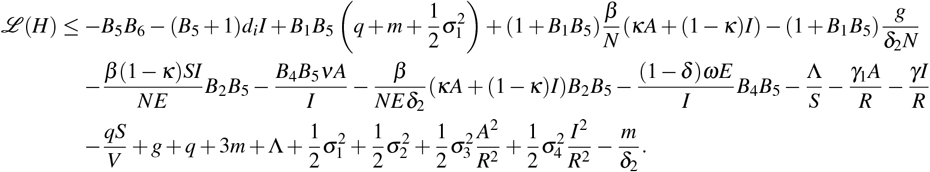

By selecting sufficiently small value of *δ*_2_ > 0, we can attain that *ℒ*(*H*) *<* 0 for every (*S, E, A, I, R,V*)∈ *U*_12_. Therefore, condition (b) of Lemma VI.1 is satisfied and hence Theorem III.2 is proved according to Lemma VI.1.

### Appendix III

One can easily write the deterministic version of the stochastic model (1) as

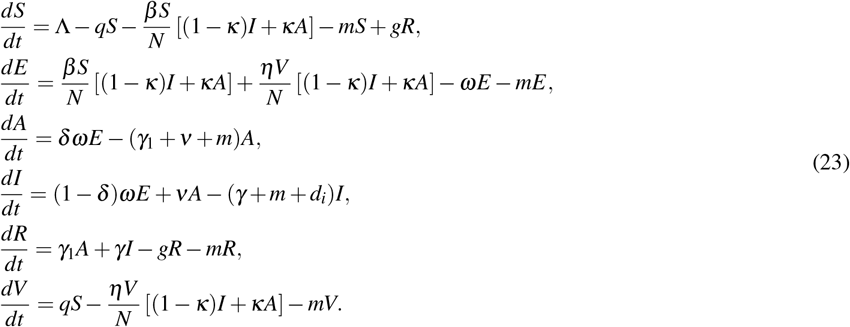

Using the next generation matrix method [41], the infection subsystem of the system (23), which describes the production of new infections and makes change in the states, reads

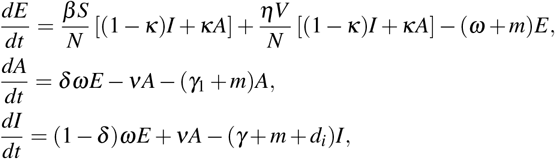

The transmission matrix (*F*) and the transition matrix (Σ) associated with the system (24) are given by

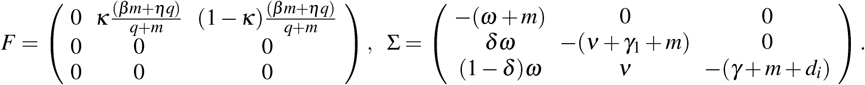

Then the deterministic basic reproduction number (DBRN) 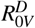 of (23) is the spectral radius of the next generation matrix −*F*Σ^−1^, i.e., 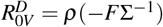, where

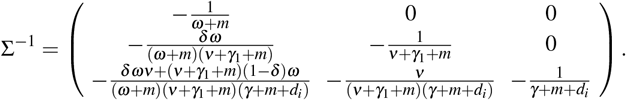

Thus,

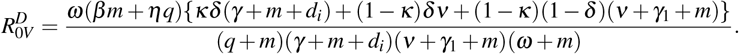

### Appendix IV

Parameter estimation has been done in two steps [32]. First, we fitted the covid data with the deterministic system (23) and next the optimal noise intensities are determined to find the best fitted parameter set for the stochastic system (II). In order to find the best fitting parameter values of the deterministic system, we used a MATLAB embedded function, lsqcurvefit, which is a nonlinear solver that minimizes the sum of squared difference between the model output and a set of data. Here a curve *y* = *f* (*t, θ*) parameterized by *θ* = (*θ*_1_, *θ*_2_, …, *θ*_*m*_) is fitted with the data points (*t*_1_, *y*_1_), (*t*_2_, *y*_2_), …(*t*_*m*_, *y*_*m*_). The non-linear least-square method finds the certain value of the parameters such that 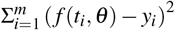 becomes minimum. With this best fit parameter set, we then find the optimum noise intensity for the stochastic system (1). Assuming 10,000 random values of *σ*_1_, *σ*_2_, *σ*_3_, *σ*_4_ between 0 to 1, the stochastic system (II) is simulated 1000 times for each of these four tuples (*σ*_1_, *σ*_2_, *σ*_3_, *σ*_4_). We then take the mean of those 1000 evolutions, and the corresponding *r*-squared value is computed. The particular value of *σ*_1_, *σ*_2_, *σ*_3_, *σ*_4_ for which the *r*-squared value is closest to 1 is our required noise intensity.

## REFERENCE

[1] E. Pritchard, P. C. Matthews, N. Stoesser, D. W. Eyre, O. Gethings, K.-D. Vihta, J. Jones, T. House, H. VanSteenHouse, I. Bell, et al., Nature Medicine, 1 (2021).

[2] WHO, in COVID-19 vaccines WHO EUL issued (WHO, 2021) pp. https://extranet.who.int/pqweb/vaccines/vaccinescovid–19–vaccine–eul–issued.

[3] O. W. in Data, in ourworldindata.org/covid-vaccinations?country=OWID WRL (2020).

[4] H. Rossman, S. Shilo, T. Meir, M. Gorfine, U. Shalit, and E. Segal, Nature medicine, 1 (2021).

[5] A. Roghani, JMIRx Med 2(3):e29324 (2021).

[6] WHO, in Global research and innovation forum (2020).

[7] K. Prem, Y. Liu, T. W. Russell, A. J. Kucharski, R. M. Eggo, N. Davies, S. Flasche, S. Clifford, C. A. Pearson, J. D. Munday, et al., The Lancet Public Health 5, e261 (2020).

[8] A. Paul, S. Chatterjee, and N. Bairagi, Medrxiv (2020).

[9] S. Moore, E. M. Hill, M. J. Tildesley, L. Dyson, and M. J. Keeling, The Lancet Infectious Diseases 21, 793 (2021).

[10] S. Khajanchi and K. Sarkar, Chaos: An Interdisciplinary Journal of Nonlinear Science 30, 071101 (2020).

[11] C. Mondal, D. Adak, A. Majumder, and N. Bairagi, ISA Transactions (2020).

[12] K. Sarkar, S. Khajanchi, and J. J. Nieto, Chaos, Solitons & Fractals 139, 110049 (2020).

[13] C. Anastassopoulou, L. Russo, A. Tsakris, and C. Siettos, PloS one 15, e0230405 (2020).

[14] M. Perc, N. Gorišek Miksić, M. Slavinec, and A. Stožer, Frontiers in Physics 8, 127 (2020).

[15] A. Majumder, D. Adak, and N. Bairagi, Stochastic Analysis and Applications, 1 (2021).

[16] K. Karako, P. Song, Y. Chen, and W. Tang, Bioscience Trends (2020).

[17] Y. Zhang, C. You, Z. Cai, J. Sun, W. Hu, and X.-H. Zhou, MedRxiv (2020).

[18] L. E. de Sousa, P. H. de Oliveira Neto, and D. A. da Silva Filho, Physical Review E 102, 032133 (2020).

[19] D. Adak, A. Majumder, and N. Bairagi, Chaos, Solitons & Fractals 142, 110381 (2021).

[20] M. R. Musa and S. Iyaniwura, MedRxiv (2021).

[21] M. De la Sen and A. Ibeas, Advances in Difference Equations 2021, 1 (2021).

[22] Y. Furuse, Journal of Global Health 11 (2021).

[23] C. Merow and M. C. Urban, Proceedings of the National Academy of Sciences 117, 27456 (2020).

[24] K. Desmet and R. Wacziarg, Journal of Urban Economics, 103332 (2021).

[25] A. Sanyaolu, C. Okorie, A. Marinkovic, R. Patidar, K. Younis, P. Desai, Z. Hosein, I. Padda, J. Mangat, and M. Altaf, SN Comprehensive Clinical Medicine, 1 (2020).

[26] J. A. Juno and A. K. Wheatley, Nature Medicine 27, 1874 (2021).

[27] J. Croda and O. T. Ranzani, The Lancet Infectious Diseases (2021).

[28] C. F. Manski and F. Molinari, Journal of Econometrics 220, 181 (2021).

[29] H. Bhapkar, P. N. Mahalle, N. Dey, and K. Santosh, Journal of Medical Systems 44, 1 (2020).

[30] E. Dolgin et al., Nature 597, 606 (2021).

[31] D. Zhou, M. Liu, and Z. Liu, Advances in Difference Equations 2020, 1 (2020).

[32] A. Majumder, D. Adak, and N. Bairagi, Physical Review E 103, 032412 (2021).

[33] Q. Yang and X. Mao, Mathematical Biosciences and Engineering 11, 1003 (2014).

[34] A. Majumder, D. Adak, and N. Bairagi, Applied Mathematical Modelling 89, 1382 (2021).

[35] C. Chen and Y. Kang, Applied Mathematical Modelling 40, 6051 (2016).

[36] V. V. Petrov, Theory of Probability & Its Applications 14, 183 (1969).

[37] O. P. Choudhary, P. Choudhary, and I. Singh, The Lancet Infectious Diseases 21, 1483 (2021).

[38] X. Mao, Stochastic Differential Equations and Applications (Elsevier, 2007).

[39] Y. Zhou, W. Zhang, and S. Yuan, Applied Mathematics and Computation 244, 118 (2014).

[40] Q. Han, L. Chen, and D. Jiang, Scientific reports 7, 1 (2017).

[41] O. Diekmann, J. Heesterbeek, and M. G. Roberts, Journal of the Royal Society Interface 7, 873 (2010).

